# The COVID-19 pandemic in Canada’s provinces: mitigation measures and outcomes

**DOI:** 10.1101/2024.06.28.24309653

**Authors:** Paul Grootendorst

**Affiliations:** Leslie Dan Faculty of Pharmacy University of Toronto Toronto ON, Canada; Department of Economics McMaster University Hamilton ON, Canada

## Abstract

The rapid spread of SARS-CoV-2 in early 2020 forced provincial health authorities across Canada to quickly institute infection control measures. It is now four years since the start of the global pandemic, and an opportune time to consider how Canada’s provinces compared in their SARS-CoV-2 containment policies and the resulting impacts on mortality and economic activity. I compare provincial exposure to SARS-CoV-2 using data on the number out-of-province arrivals into each province. I compare the key containment measures used in each province, the length of time that these measures were imposed, and the uptake of COVID-19 vaccines by province. Using Statistics Canada data, I also estimate the impact of the COVID-19 pandemic on provincial crude death rates during 2020-2023, and life expectancy and gross domestic product during 2020-2022. I find substantial provincial variation in pandemic responses and outcomes. The provinces varied in their use of the most stringent public health measures. Uptake of the primary COVID vaccinations varied from 76% to 92%; booster vaccination uptake varied even more. There was also marked provincial variation in mortality and economic outcomes. While this study does not estimate the impacts of provincial response stringency and COVID vaccine uptake on mortality and other outcomes, it does provide suggestive evidence that can be formally assessed in future research.

## Introduction

The rapid spread of the severe acute respiratory syndrome coronavirus 2 (SARS-CoV-2) in early 2020 challenged governments and public health authorities in Canada and globally. The first known diagnosis of SARS-CoV-2 infection in Canada was made on January 25, 2020.[1,2] Less than seven weeks later, on March 11, the World Health Organization (WHO) declared the novel coronavirus disease (COVID-19) outbreak a global pandemic.[3] (COVID-19 is the name of the disease caused by SARS-CoV-2.) Authorities in each province and territory quickly decided how to best contain the spread of SARS-CoV-2 in their jurisdictions using school and business closures and other mitigation measures;[4] the federal government, meanwhile, was responsible for international border controls,[5] and procuring testing kits, face masks, and other protective equipment.

Four years after the WHO declared the global pandemic, it is now an opportune time to consider how Canada’s provinces compared in their SARS-CoV-2 containment policies and the resulting impacts on mortality and economic activity. Data on the number of international and domestic arrivals into each province is used to compare their initial exposure to SARS-CoV-2. I assess key containment measures used in each province, the length of time that these measures were imposed, and the uptake of COVID-19 vaccines after these became widely available in early 2021. Using newly available data, I also estimate the impact of the COVID-19 pandemic on provincial crude death rates, life expectancy and gross domestic product. The paper builds on the work of McGrail [6] who reported province-level COVID excess mortality (estimated by Statistics Canada) and COVID-attributed deaths during the first 19 months of the pandemic. I do not attempt to formally link outcomes to public health mitigation measures and vaccine uptake, although the data presented here provides some suggestive evidence.

## Methods

### Travel patterns

As SARS-CoV-2 originated outside of Canada, the provinces that were the most common destinations of trips originating outside of Canada were, at least initially, most susceptible to viral spread. To assess this risk, I used Canada Border Services Agency data, compiled by Statistics Canada, on the number of individuals (both residents of Canada and non-residents) entering Canada by air, land or water.[7] These data are stratified by the province and month of entry. I use Statistics Canada’s National Travel Survey (NTS) trip file, [8] to assess the volume of travel within Canada. Each year, the NTS samples Canadian households and within households, a household member to provide basic information, including the trip destination, on all his or her trips (domestic and international) that ended in the reference month. Sampling weights are provided to render estimates representative of the residential population. I use the NTS trip file data to determine, by quarter, the number of interprovincial trips, by province of destination and quarter.

### Non-pharmaceutical containment measures

The Oxford Coronavirus Government Response Tracker (OxCGRT) group collected information on nine key policy responses that governments in 185 countries have taken to increase social distancing during the pandemic.[9] These policy responses are school closures; workplace closures; cancellation of public events; restrictions on public gatherings; closures of public transport; stay-at-home requirements; public information campaigns; restrictions on internal movements; and international travel controls. OxCGRT gathered information on each type of policy response daily from January 2020 to December 2022. Responses were measured on an ordinal scale with higher values representing higher levels of restriction. For instance, the stay-at-home response levels are: 0 (no measures), 1 (a public advisory to stay home), 2 (a stay-at-home mandate with exceptions for “daily exercise, grocery shopping, and ‘essential’ trips”) and 3 (a mandate with only very limited exceptions). If the restrictions were not generally applied, but were instead targeted by geography or along other dimensions, then response levels were adjusted downwards. The nine adjusted responses were aggregated to create a daily government “stringency index”. [10]

Analysts at the Bank of Canada (BOC) adapted the OxCGRT stringency index to capture provincial policy responses each day from January 1, 2020 to July 19, 2022. [11] The BOC and OxCGRT stringency indices differ in several ways. First, the BOC index replaces the OxCGRT “restrictions on internal movements” response with two responses capturing separately restrictions on intra-provincial travel and inter-provincial travel. The BOC index also adds two policy responses not found in the OxCGRT index; these capture separate enforcement mechanisms for individuals and for firms violating public health orders. The BOC index adds more targeting dimensions than the OxCGRT index; for example, the “cancellation of public events” response is adjusted downwards if only indoor events are cancelled. Finally, the BOC adjusted the number of levels for some policy responses. [11] Each of the 12 policy responses were measured on a scale from 0, no restrictions, to 100, representing the highest level with no targeting adjustments. The overall BOC stringency index is a simple average of the 12 subindices.

I used these data to graph, by province, the weekly average BOC stringency index, and the number of new COVID cases per 100,000 reported by the Public Health Agency of Canada (PHAC) during 2020-2022.[12] These graphs reveal how provincial policy makers adjusted stringency levels in response to changes in COVID infection. I also identify, for each province, the timing and duration of 5 particularly stringent policy responses. These include: 1) a stay-at-home order index value of 66 or more, where 66 reflects a province-wide stay home order with exceptions allowed for daily exercise, grocery shopping and essential trips; 2) a school and university closure index value of 58 or more, where 58 reflects closure of some but not all types of institutions (such as elementary schools but not universities) with an allowance made for on-line learning; 3) a workplace closure index of 66 or more, where 66 reflects closing (or work from home) for all but a broad set of essential workplaces, albeit with an allowance for business curbside pickups; 4) a restrictions on private gatherings index of 80 or more, where 80 reflects a restriction limit on gatherings of 15 people or less; and 5) a restrictions on interprovincial travel index value of 55 or more, where 55 reflects a quarantine requirement for interprovincial travellers with exceptions given to arrivals from designated provinces. I graphed for each province the weeks during which each of these types of restrictions were in effect, and superimposed the number of new COVID deaths per 100,000 reported by PHAC. The 7 other policy responses contained in the BOC stringency index, such as the cancelling of public events, and information campaigns, tend to be implied by the policy responses I focus on.

### Vaccination campaigns

I used data from PHAC [13] to graph, by province, the percentage of the population that completed their primary COVID vaccination series by week from January 2021 (the first full month of vaccine rollout) to June 2022. I graph vaccine uptake separately by sex (male, female) and again by age group (age 18-29, 40-49, 60-69, 80+). Most of the vaccines used in Canada require two doses; the Janssen Ad26.COV2.S is the sole single-shot vaccine. I also graph PHAC data on the percentage of the population that received their first and second COVID booster vaccinations by sex and week from July 2021 to June 2023.

### Economic outcomes

I graphed, for each province, Statistics Canada annual data on real (i.e. inflation-adjusted) provincial gross domestic product (GDP), measured in 2017 dollars.[14] GDP was expressed in per capita terms using the Statistics Canada population data described earlier. To estimate the impact of COVID on GDP, I extrapolated the trend line in GDP during the 5 pre-pandemic years, 2015-2019, into the pandemic period, 2020-2022, and compared to actual GDP during this period. I estimated the changes for each year, along with their 95% confidence intervals.

### Mortality outcomes

Next, I graphed province-specific excess mortality rates as estimated by Statistics Canada.[15] These estimates are provided weekly from the January 2020 to August 2023. Note that this “excess mortality” approach captures the net effect of the pandemic. This is the mortality from SARS-CoV-2 infection plus any deleterious effects of lockdowns, such as delayed access to healthcare [16] less any offsets due to the COVID era lockdowns and other social distancing measures, such as a reduction in road traffic collisions [17] and the reduced mortality from seasonal influenza and other infectious disease. I also present the crude all-cause mortality rate, by sex, province and week during the years 2010-2023 that Statistics Canada used to estimate excess mortality. To calculate mortality rates, I used weekly death counts and annual population counts, specific for each sex and province; these data were collected by Statistics Canada.[18,19]

To assess the impact of COVID-19 on age-specific mortality, I used Statistics Canada’s annual estimates of life expectancy at birth and age 65, by sex and province. This statistic estimates, for each year, the number of years an individual of a specific age (in this case 0 or 65) can expect to live if they face in future years the same annual mortality probabilities that older individuals – of the same sex and province – experienced during the year. [20] (These mortality probabilities are obtained from Statistics Canada’s period life tables. [21]) Life expectancy at 65 during a pandemic year thus reflects the annual mortality probabilities of those aged 65 or older in the same year. Statistics Canada did not release life expectancy values for Prince Edward Island because the province’s small population renders the values highly variable. To estimate the impact of COVID on life expectancy, I used the same extrapolation approach as was used for real per capita GDP. Analyses were conducted using Stata v17.[22]

## Results

### Travel patterns

Fig 1 presents data on the number of individuals entering Canada, by province of entry and month. Prior to the pandemic, Ontario, British Columbia and Quebec received by far the largest number of arrivals. The border restrictions imposed by the federal government drastically reduced arrivals in the first months of the pandemic. Despite these restrictions, however, 353,000 individuals entered Ontario from abroad in April 2020, the first full month of the pandemic. Arrivals into British Columbia and Quebec in the same month were 94,000 and 70,000. Arrivals into the other provinces were much smaller. Fig 2 displays the total of international arrivals and arrivals from other provinces, by province of arrival and quarter. Ontario, British Columbia, Alberta and Quebec received the largest number of arrivals; arrivals into the other provinces were markedly lower.

**Fig 1.**
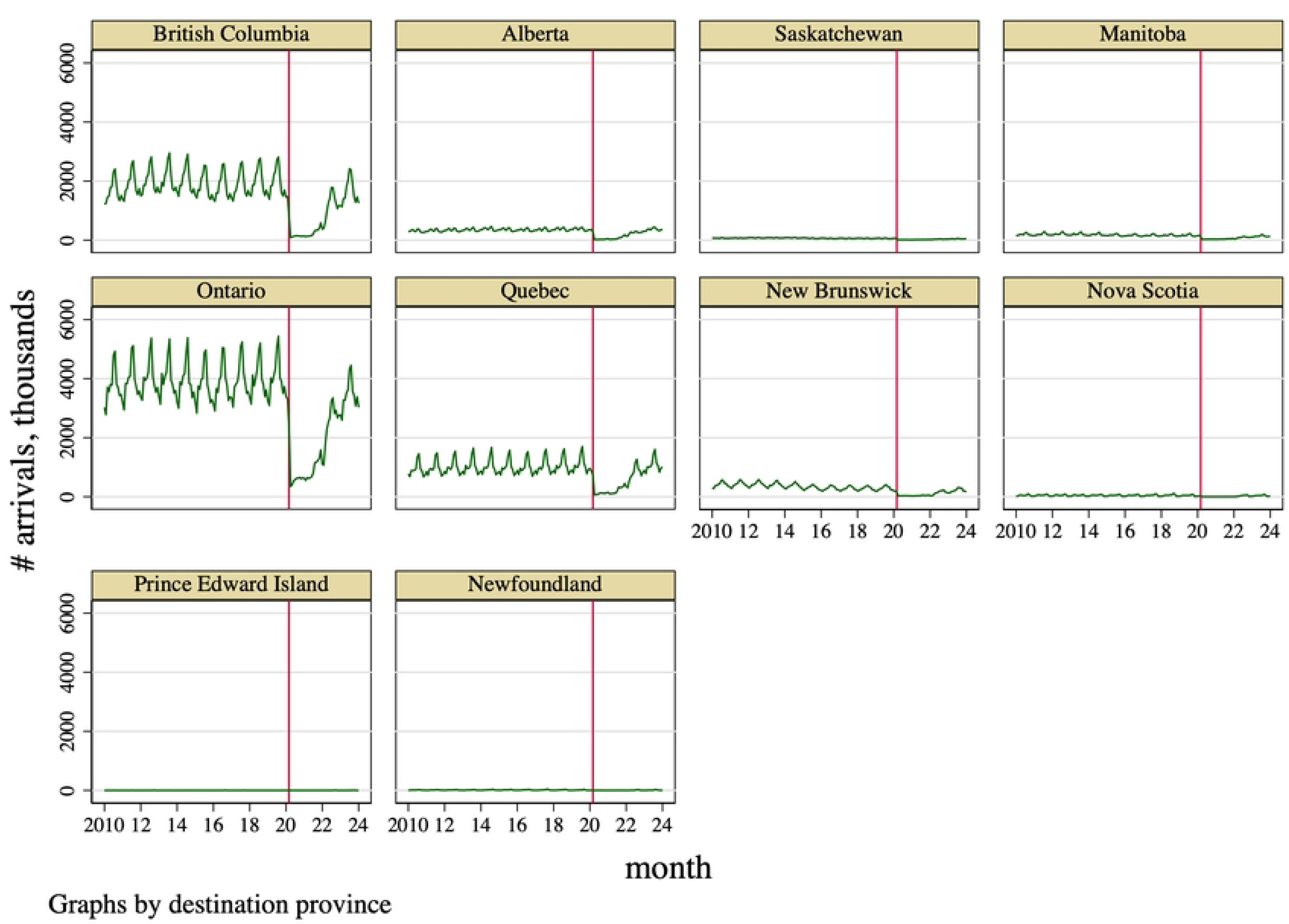
Number of international arrivals, by province and month of arrival, 2010m1 to 2024m1. Data source: Statistics Canada. Table 24-10-0053-01 International travellers entering or returning to Canada, by type of transportation and traveller type. Vertical line indicates the month that the WHO declared the global COVID-19 pandemic.

**Fig 2.**
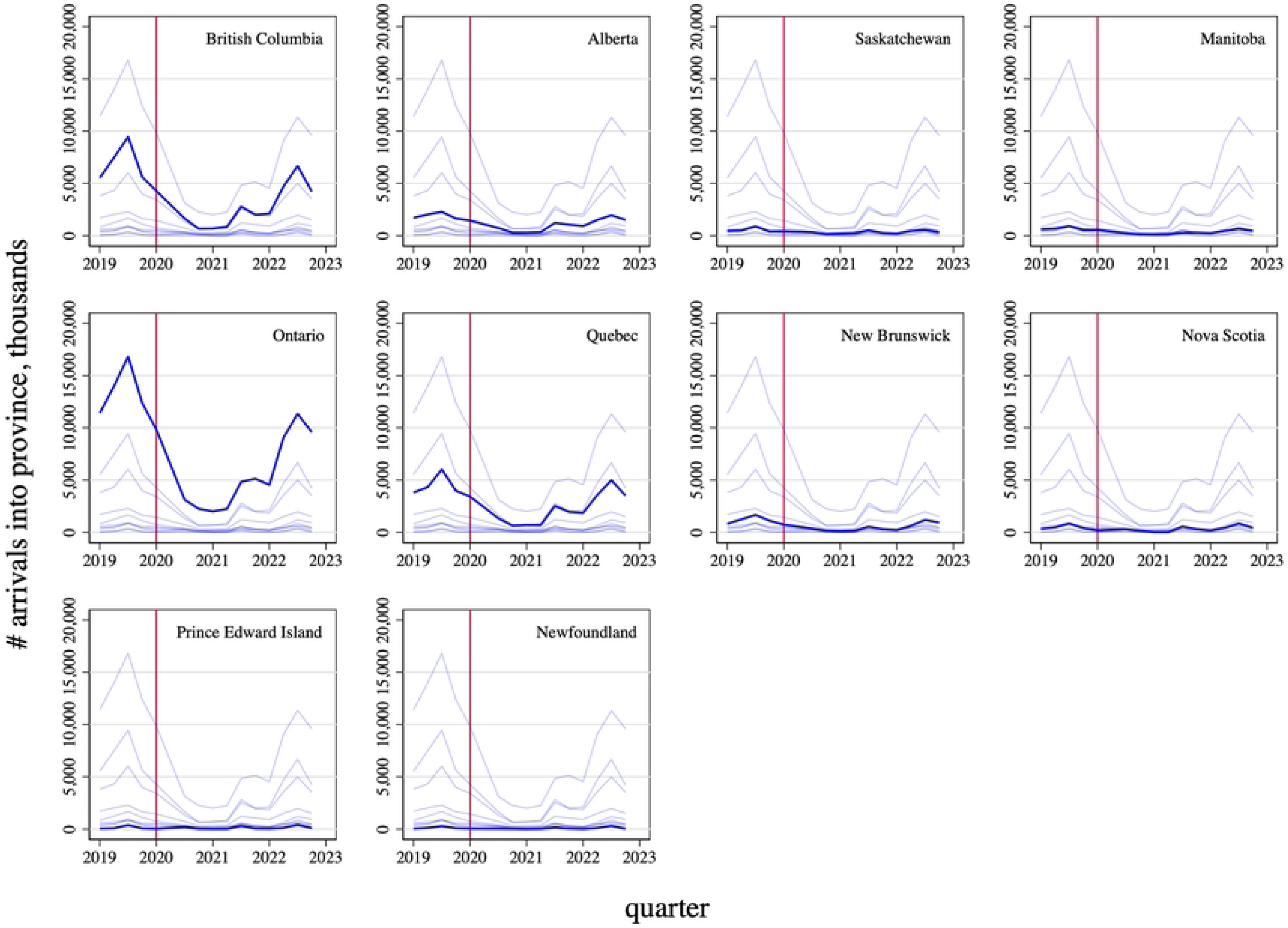
Total of international arrivals and arrivals from other provinces, by province and quarter of arrival, 2019q1 to 2022q4. Data source: International arrivals: Statistics Canada. Table 24-10-0053-01 International travellers entering or returning to Canada, by type of transportation and traveller type. Domestic arrivals: Statistics Canada National Travel Survey. Vertical line indicates the quarter that the WHO declared the global COVID-19 pandemic.

### Overall stringency of provincial public health measures

The provincial stringency indices, displayed in Fig 3, varied with the different waves of the pandemic; these waves are illustrated by the periodic increases in new COVID cases per 100,000. The provinces tended to impose the most stringent controls at the start of a wave of infection, especially the first wave in March 2020, and then ease these restrictions until the next wave occurred, at which point restrictions were again tightened.[5,23–25] The degree of restriction at the start of a wave, however, tended to decline after the summer of 2021, about 6 months into the COVID vaccination campaign. By July 2022, most restrictions were eased. The provinces varied in the degree of restriction across the pandemic. The three westernmost provinces tended to have lower stringency index values at a given date, especially after the first wave, compared to the other provinces.

**Fig 3.**
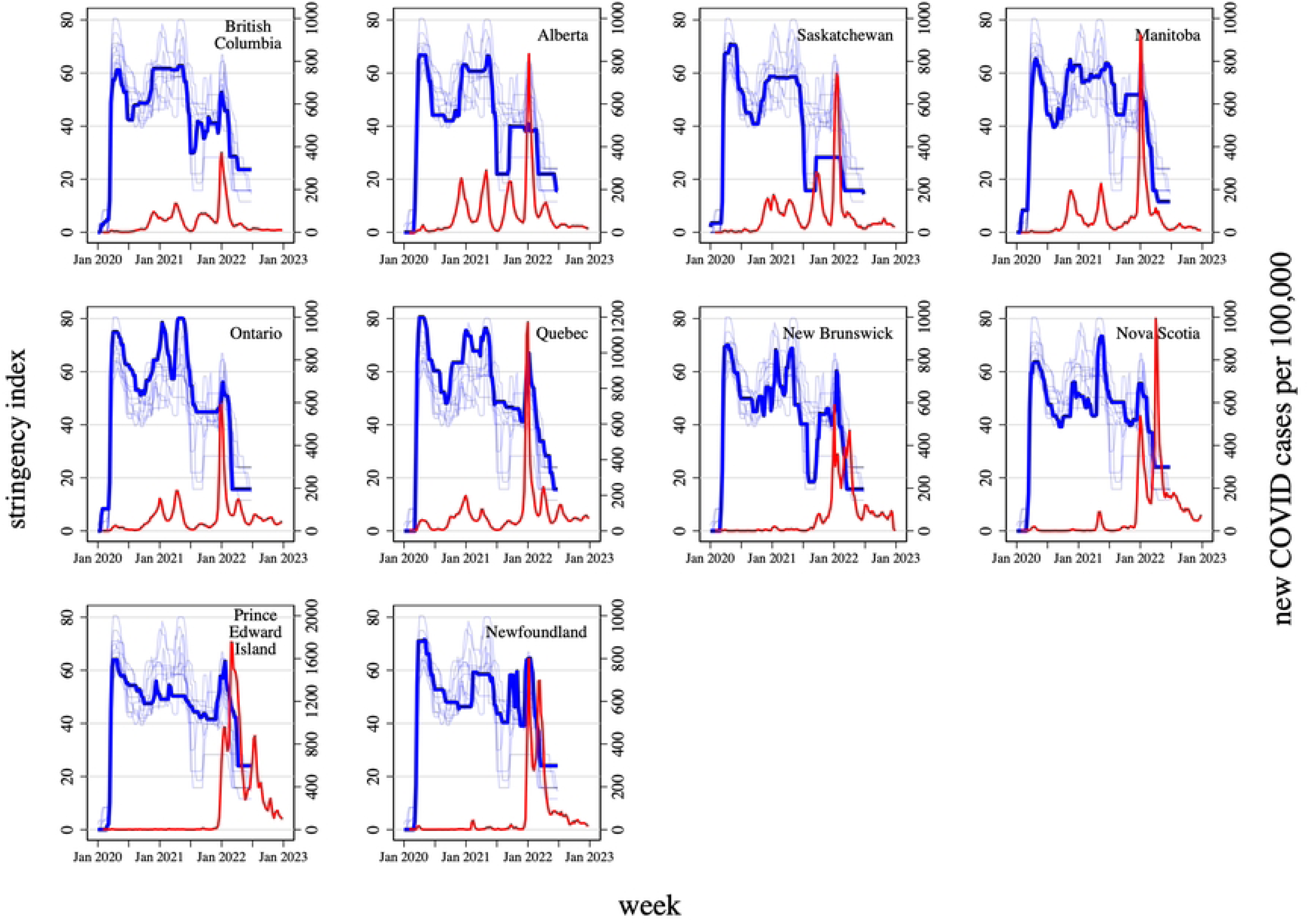
Bank of Canada stringency index values, by province and week, January 2020 - July 2022 and new COVID cases per 100,000, by province and week, January 2020 - December 2022. Data source: Bank of Canada; Public Health Agency of Canada COVID-19 epidemiology update: Summary. Bank of Canada stringency index (left hand side y-axis) in blue; new COVID-19 cases per 100,000 population (right y-axis) in red. Each graph plots stringency indices for each province, but bolds the index for just one province. Note the scale of COVID cases (right y-axis) varies by province.

Fig 4 provides insights into the timing and duration of the most restrictive provincial infection mitigation measures: school and workplace closures (respectively, shaded orange with dashed border, and shaded green with long-dash border), stay-at-home orders (shaded blue with a solid border), 15 person or less limit on gatherings (shaded gray with dotted border), and quarantine requirements for domestic arrivals from other provinces (shaded yellow). I call these “type 1 restrictions” for brevity. The figure highlights the weeks during which each of these type 1 restrictions were in effect; the COVID-attributed mortality rate is superimposed.

**Fig 4.**
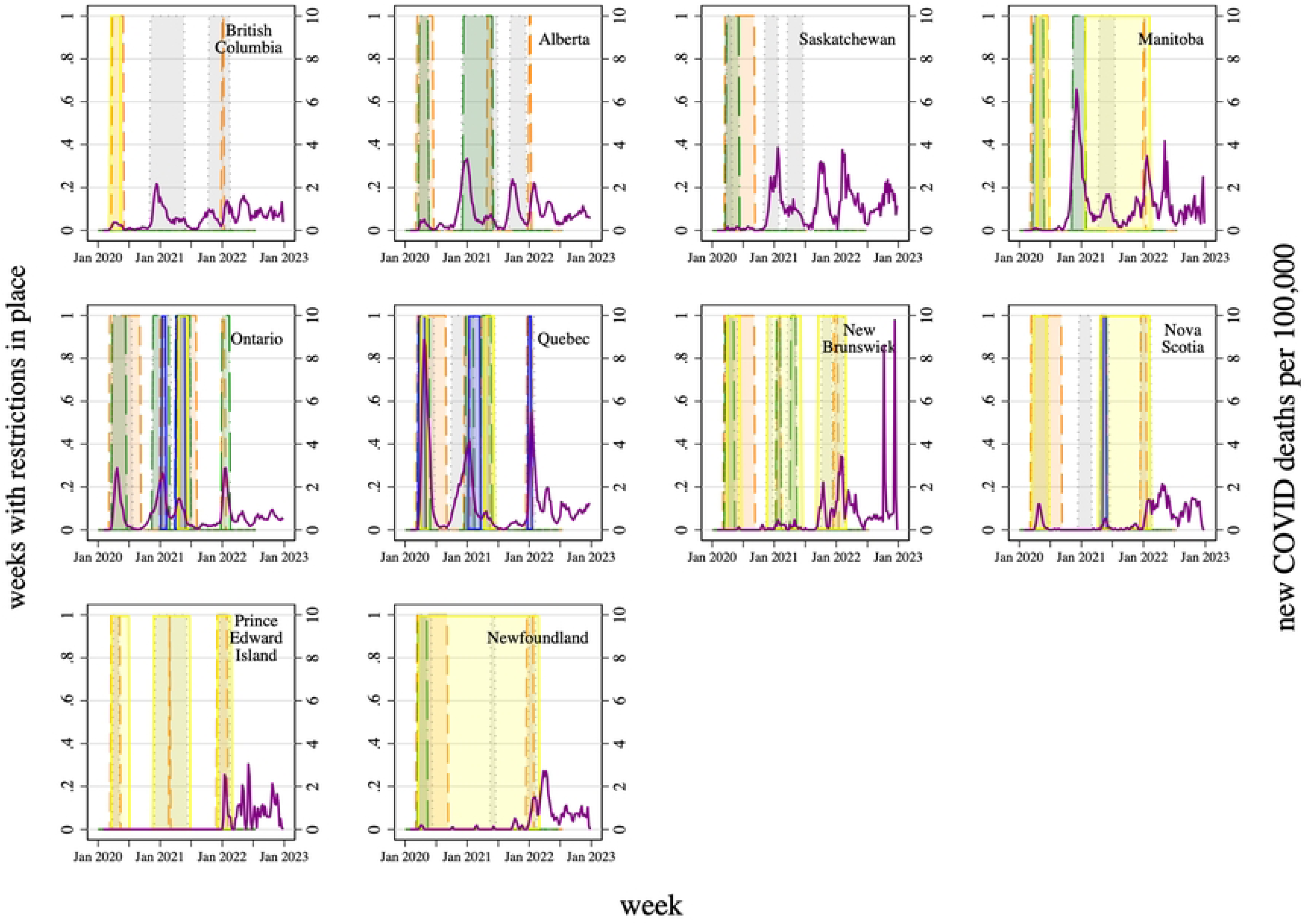
Weeks with social distancing restrictions in effect, by restriction type, province, and week, January 2020 - July 2022, and new COVID-attributed deaths per 100,000 population, by province, and week, January 2020 - December 2022. Data source: Bank of Canada; Public Health Agency of Canada. Left hand side y-axis reflect weeks during which restriction levels greater than or equal to a minimum level of stringency were in place. Gray with dotted border: restriction limit on private gatherings of 15 people or less. Orange with dashed border: some educational institutions closed. Green with long-dash border: closing (or work from home) for all but a broad set of essential workplaces, with an exception for curbside pickups. Blue with solid border: stay at home order with some exceptions for daily exercise, grocery shopping, and ‘essential’ trips. Yellow: quarantine requirement for interprovincial travellers with exceptions given to arrivals from designated provinces. New deaths attributed to COVID-19 per 100,000 population (right hand side y-axis) represented by the purple line.

The four provinces in the Atlantic region – New Brunswick, Nova Scotia, Prince Edward Island and Newfoundland – had low numbers of arrivals from outside of this region during the first quarter of 2020. Nevertheless, authorities used school and workplace closures that extended until the fall of 2020 to limit infection during the first wave of the pandemic. The region also used border controls that extended up to 22 months into the pandemic; those that entered the region were required to self-isolate.[26] Indeed, travel into the Atlantic region was so low during the pandemic (Fig 2) that airlines discontinued service to the area. [27] With strict border controls in place, the four Atlantic provinces surgically applied type 1 restrictions to reduce infection during subsequent waves. This strategy kept COVID infection and mortality in the Atlantic region very low, at least until border controls and other type 1 restrictions were removed in March 2022. After this, COVID-attributed mortality rates grew markedly. Indeed, New Brunswick experienced several weeks with 10 COVID-attributed deaths per 100,000 in the fall of 2022.

The pandemic evolved differently in other provinces. In the first quarter of 2020, Ontario and Quebec were common destinations of trips originating outside of Canada. Perhaps because of the inflow of individuals infected with SARS-CoV-2, Ontario and especially Quebec experienced large surges in mortality during the first wave of the pandemic despite school and workplace closures. These stringent restrictions were eased after the first wave but were quickly reinstated as mortality again grew in the late fall of 2020. Authorities in these provinces used type 1 policies, including stay-at-home orders, for relatively long periods of time to mitigate the spread of infection in subsequent waves of the pandemic. Mortality rates were relatively low by the start of summer of 2021, resulting in an easing of restrictions, but infection and mortality again increased in the fall of 2021, causing restrictions to be tightened over the holiday season. Mortality rates subsequently declined over the winter of 2022.

Manitoba was able to control the first wave with school and workplace closures and quarantine requirements for visitors. These restrictions were relaxed by July 2020. But mortality rates increased markedly in the late fall 2020, prompting authorities to again impose workplace closures until mortality rates dropped, in February 2021. Thereafter, Manitoba re-imposed quarantine requirements for visitors, and in the face of an increase in COVID infection and mortality in the spring of 2021, imposed a restriction on the size of private gatherings. All type 1 restrictions were removed in early 2022; after this time, the province experienced a marked increase in COVID attributed mortality.

Alberta and Saskatchewan successfully limited first wave mortality using school and workplace closures. These restrictions were relaxed by June 2020 in Alberta and by September 2020 in Saskatchewan. Infection rates and mortality increased during the fall, causing Alberta to reintroduce strict limits on gatherings and workplace closures that extended until June 2021. Saskatchewan responded to the fall 2021 infection surge by briefly restricting private gatherings in the spring of 2021. Thereafter, neither province used type 1 restrictions to quell several subsequent waves of COVID infection.

British Columbia, despite the flow of international arrivals from Asia and elsewhere in the first months of 2020, successfully navigated the first wave of the pandemic using school closures and quarantine requirements for out of province visitors; these restrictions were in effect for just 10 weeks. Indeed, compared to Alberta and Saskatchewan, British Columbia used type 1 restrictions more sparingly, and for shorter durations; overall policy stringency, however, was higher in British Columbia. Infection rates and mortality increased during the fall and these were met with strict limits on private gatherings that were in effect until spring of 2021. Infection rates and mortality increased again in the fall of 2021, again resulting in restrictions on gatherings and a brief school closure in early 2022.

### COVID vaccine uptake

The federal government received its first COVID vaccine shipments in late 2020 and distributed these vaccines to provincial authorities who, in turn, launched mass vaccination campaigns. Overall, 81% of Canadians received their primary COVID vaccination series; most primary vaccinations had been administered within 12 months of the start of the vaccine rollout (Fig 5). The vaccinated percentage at June 30, 2022 varied by province and sex. Vaccination rates were highest in Newfoundland (around 92%) and lowest in Alberta (76%) and Saskatchewan (77%). Vaccination rates for females were about 2-3 percentage points higher than for males. Vaccination rates tended to increase with age (Fig 6). Older individuals also tended to be vaccinated first, reflecting a prioritization of vaccine access to older individuals, especially those residing in long term care facilities. [23,28]

**Fig 5.**
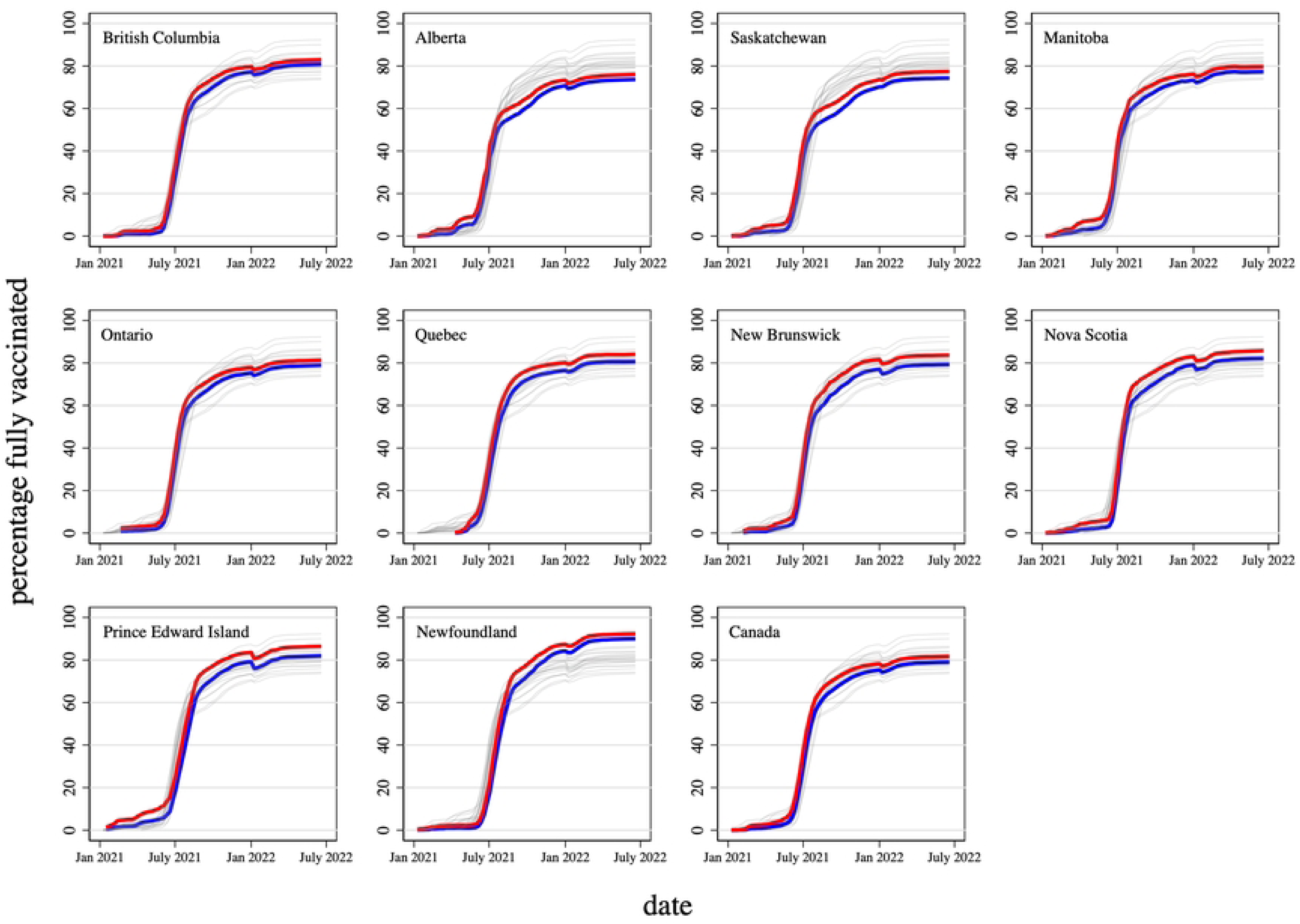
Percentage of population that completed primary vaccination series, by province, sex and date, January 1, 2021 - June 30, 2022. Data source: Public Health Agency of Canada. Canadian COVID-19 vaccination coverage report. Red line: females. Blue line: males. Each graph plots vaccine uptake for each province and both sexes, but bolds the uptake for just one province or for Canada.

**Fig 6.**
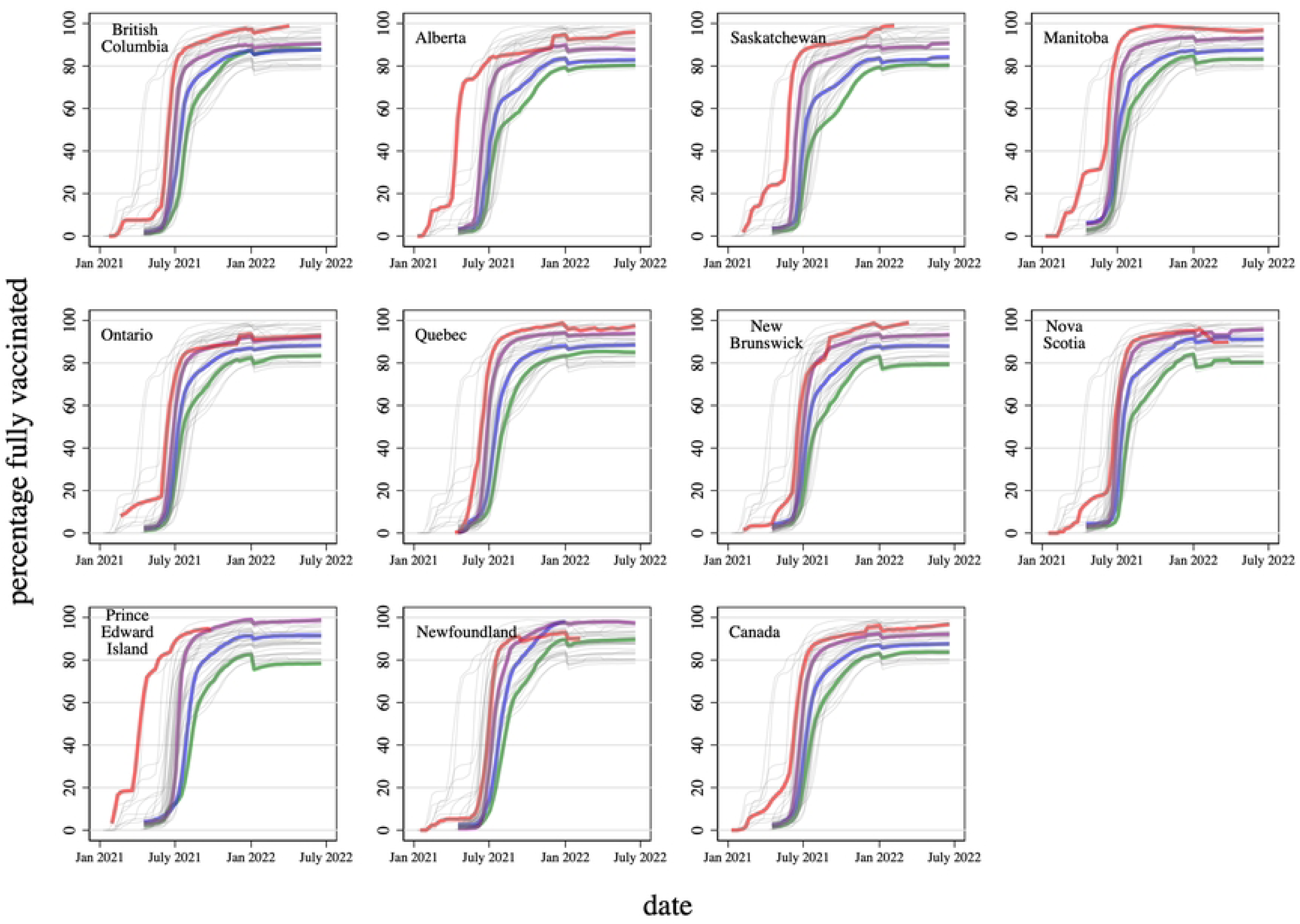
Percentage of population that completed primary vaccination series, by province, age group (18-29, 40-49, 60-69, 80+) and date, January 1, 2021 - June 30, 2022. Data source: Public Health Agency of Canada. Canadian COVID-19 vaccination coverage report. Green line: 18-29 years. Blue line: 40-49 years. Purple line: 60-69 years. Red line: 80+ years. Each graph plots vaccine uptake for each province and all four age groups, but bolds the uptake for just one province or for Canada.

About half of Canadians received the first COVID booster and about one quarter received the second COVID booster vaccine by July 2023 (Fig 7). Booster uptake among females was about 5-6 percentage points higher than for males. There was marked interprovincial variation in booster uptake; uptake was about 20 percentage points lower in Alberta than in Newfoundland.

**Fig 7.**
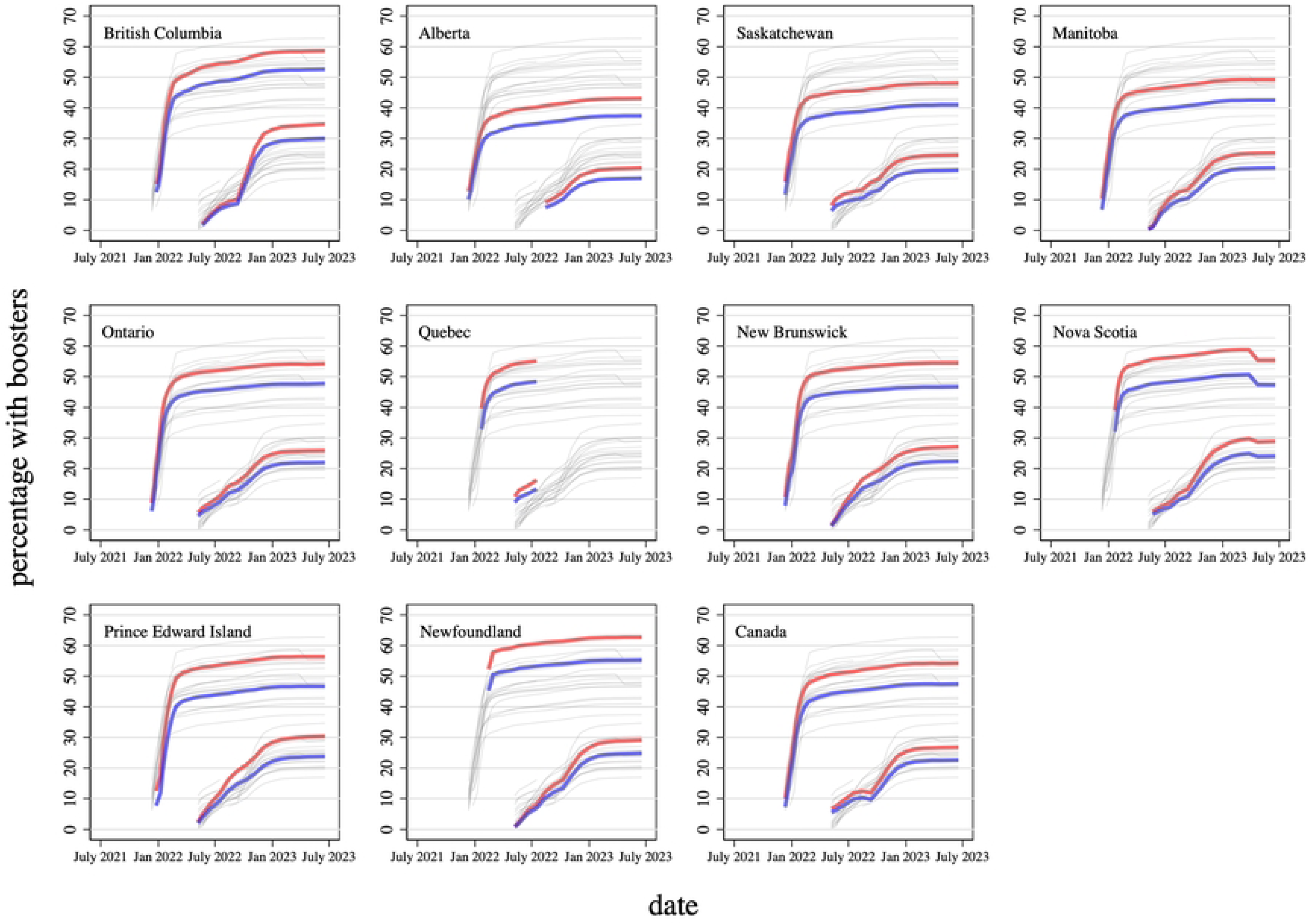
Percentage of population that received booster vaccinations, by province, booster number (first and second), sex and date, July 1, 2021 - June 30, 2023. Data source: Public Health Agency of Canada. Canadian COVID-19 vaccination coverage report. Red line: females. Blue line: males. First booster series values appear above the second booster series values. Each graph plots booster vaccine uptake for each province and each sex, but bolds the uptake for just one province or for Canada. Data for Quebec unavailable after August 2022.

### Economic activity

The lockdowns during the first three years of the pandemic inevitably reduced both productivity and the sale of goods and services. What effect did this have on real per capita GDP? In most provinces, the effects were comparable to those experienced during the 2008 global financial crisis (Fig 8). The estimated post-COVID reductions in real per capita GDP, illustrated in S1 Fig, and reported in S1 Table, varied by province and year. The average annual GDP reduction in Alberta during the period 2020-2022, estimated by comparing actual GDP each year during this period to the counterfactual obtained by extrapolating the 2015-2019 trend line, and then averaging, was $4,554 (2017 dollars). The estimated reductions in New Brunswick, Nova Scotia and Prince Edward Island were less than half this amount. Table 1 provides estimates for other provinces.

**Fig 8.**
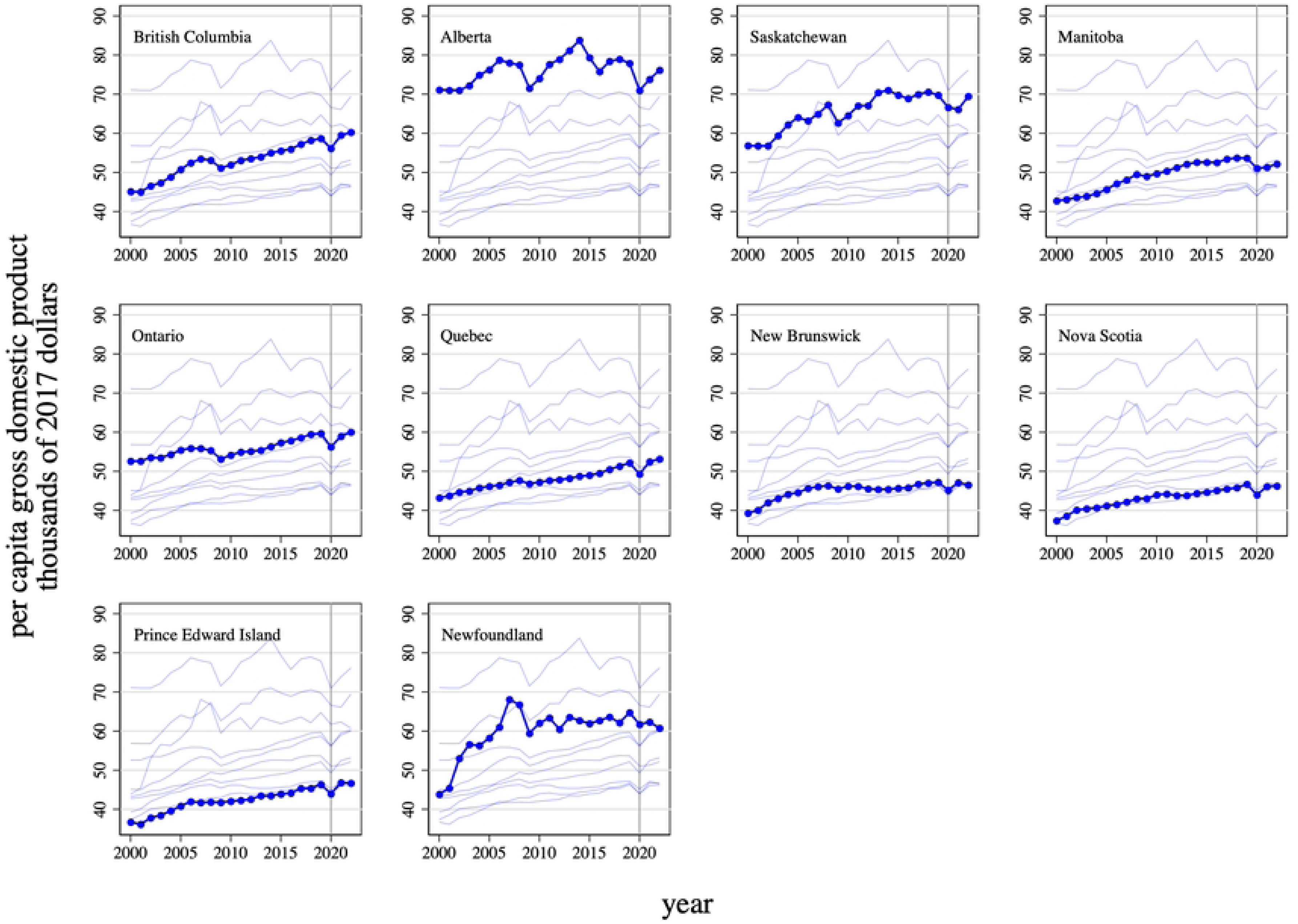
Real per capita gross domestic product, in thousands of 2017 dollars, by province and year, 2000-2022. Data source: GDP: Statistics Canada. Table 36-10-0222-01 Gross domestic product, expenditure-based, provincial and territorial, annual, measured in chained (2017) dollars. Population data: Statistics Canada. Table 17-10-0005-01 Population estimates on July 1, by age and gender. Vertical line indicates the year that WHO declared the global COVID-19 pandemic. Each graph plots real per capita GDP for each province, but bolds the real per capita GDP for just one province.

**Table 1.**
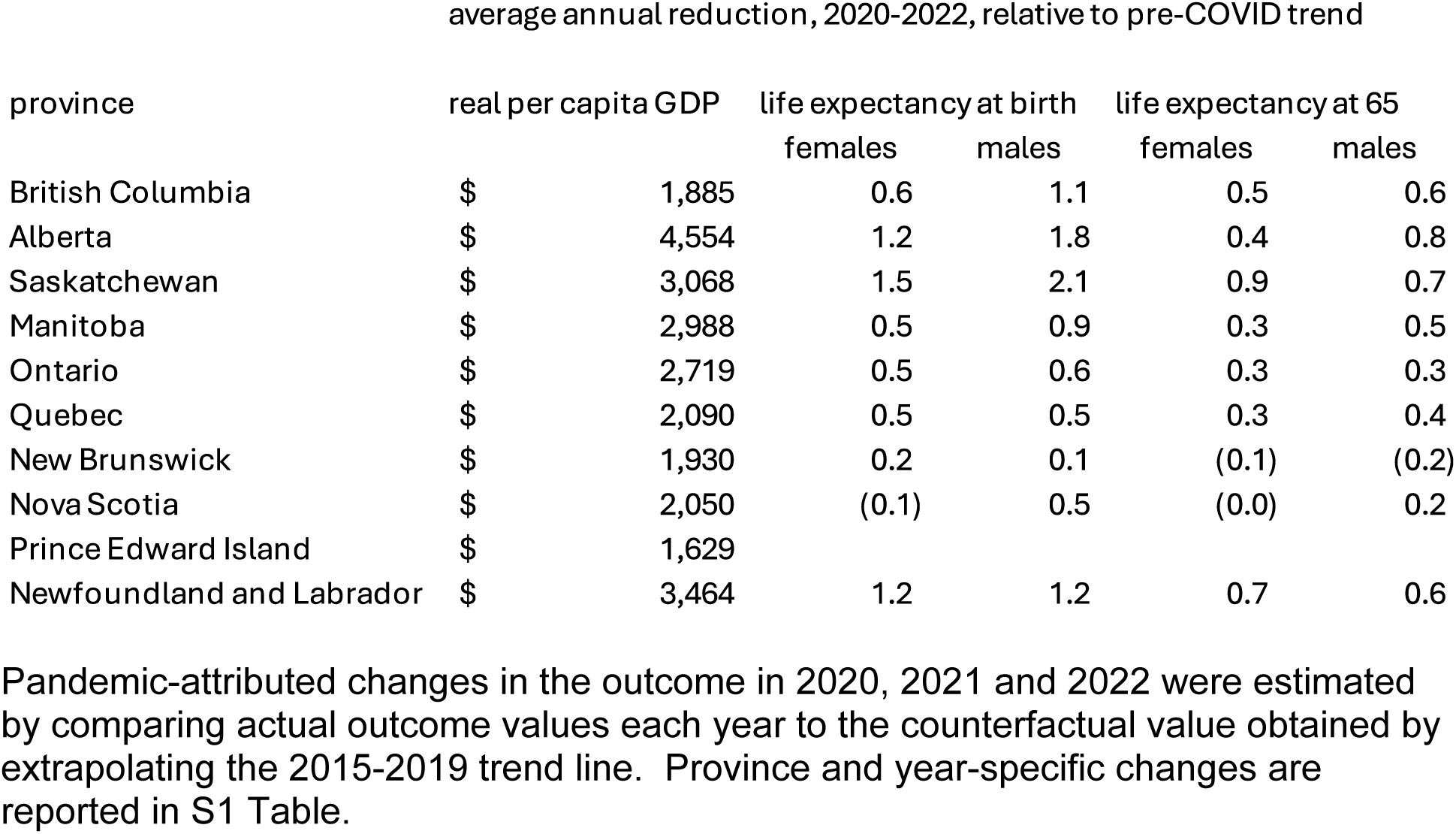
Estimated effects of the COVID-19 pandemic on real per capita gross domestic product, life expectancy at birth and life expectancy at 65.

### Mortality

Fig 9 presents Statistics Canada estimates of COVID-era excess mortality by week and province. Given the large weekly variation in provincial mortality rates, I fit median spline curves through the data; these help to illustrate seasonal and annual trends. The COVID-attributed mortality rate is superimposed, as is the rate of new COVID cases. Several patterns emerge from these data. First, there was *negative* estimated excess mortality in the Atlantic region during most of the first two years of the pandemic, when strict border controls and other social distancing measures were in place. Second, COVID-era excess mortality typically exceeds the COVID-attributed mortality reported to public health authorities. Third, estimated excess mortality tended to be highest after the most stringent social distancing measures were relaxed in July 2022. This mortality surge appears to have been caused by the immune-evasive BA.4 and BA.5 sublineages of the Omicron variant of SARS-CoV-2. [24,29] In some provinces, like New Brunswick, the post-July 2022 surges in excess mortality coincided with surges in COVID-attributed mortality reported by public health authorities. In other provinces, like Newfoundland, rates of COVID-attributed mortality were very low despite high rates of excess mortality.

**Fig 9.**
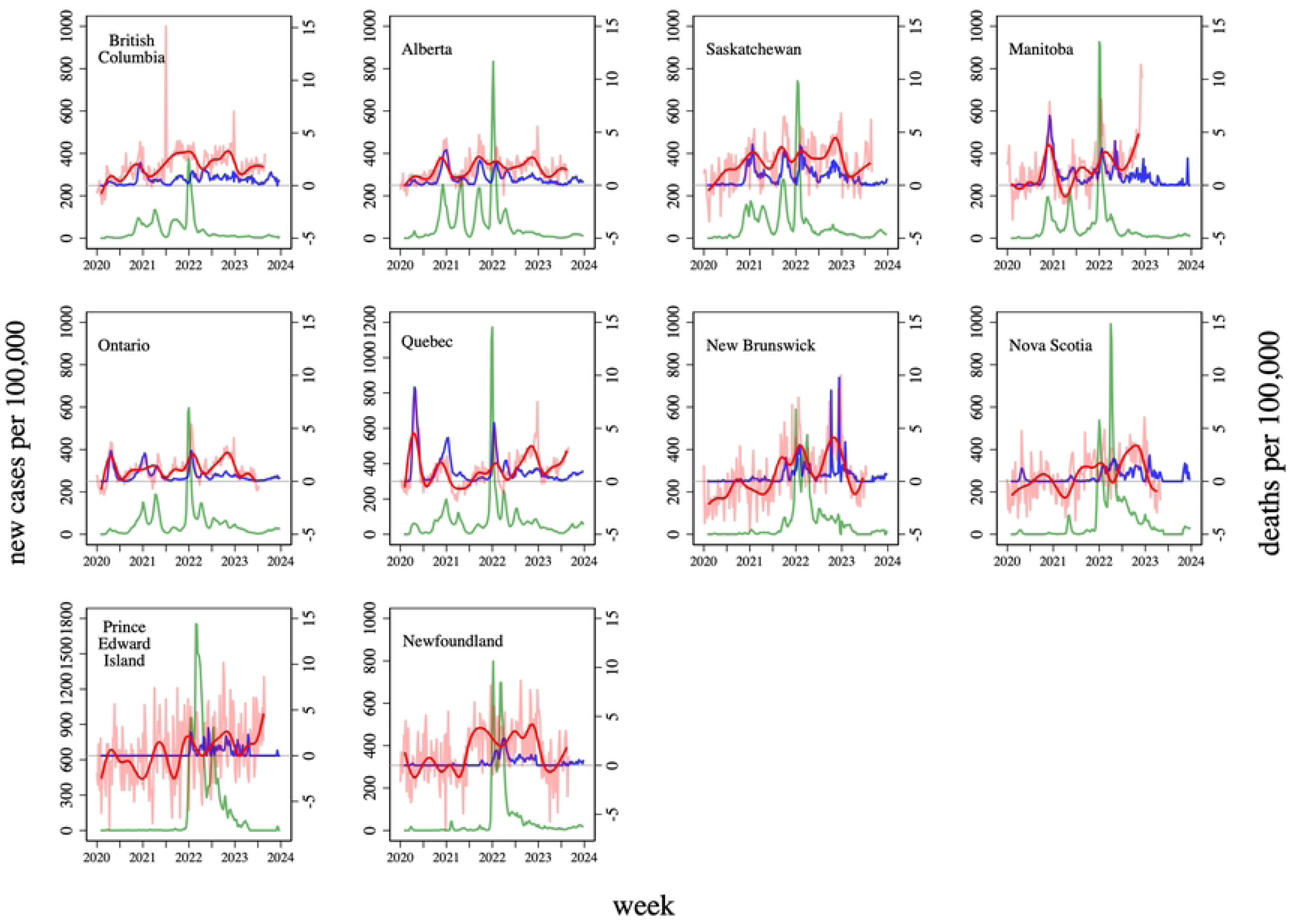
New COVID cases and COVID-attributed deaths per 100,000, by province and week, January 2020 - December 2023 and estimated excess deaths per 100,000, by province and week, January 2020 - August 2023. New COVID cases per 100,000 (left-hand side axis) in green. Estimated excess mortality per 100,000 and median spline fit to these data (right hand side axis) in red. COVID-attributed deaths per 100,000 (right-hand side axis) in blue. Data source: Public Health Agency of Canada COVID-19 epidemiology update: Summary. Estimated excess deaths: Statistics Canada. Table 13-10-0784-01 Provisional weekly estimates of the number of deaths, expected number of deaths and excess mortality, inactive. All-cause mortality rates: Statistics Canada. Table 13-10-0768-01 Provisional weekly death counts, by age group and sex. Population data: Statistics Canada. Table 17-10-0005-01 Population estimates on July 1, by age and gender.

Fig 10 displays province-level weekly trends in rates of all-cause mortality by sex over the period 2010-2023. Again, I fit median spline curves to the data given the marked weekly and seasonal variation. The figure reveals that in several provinces, notably British Columbia and Ontario, COVID-era excess mortality was higher for males than for females.

**Fig 10.**
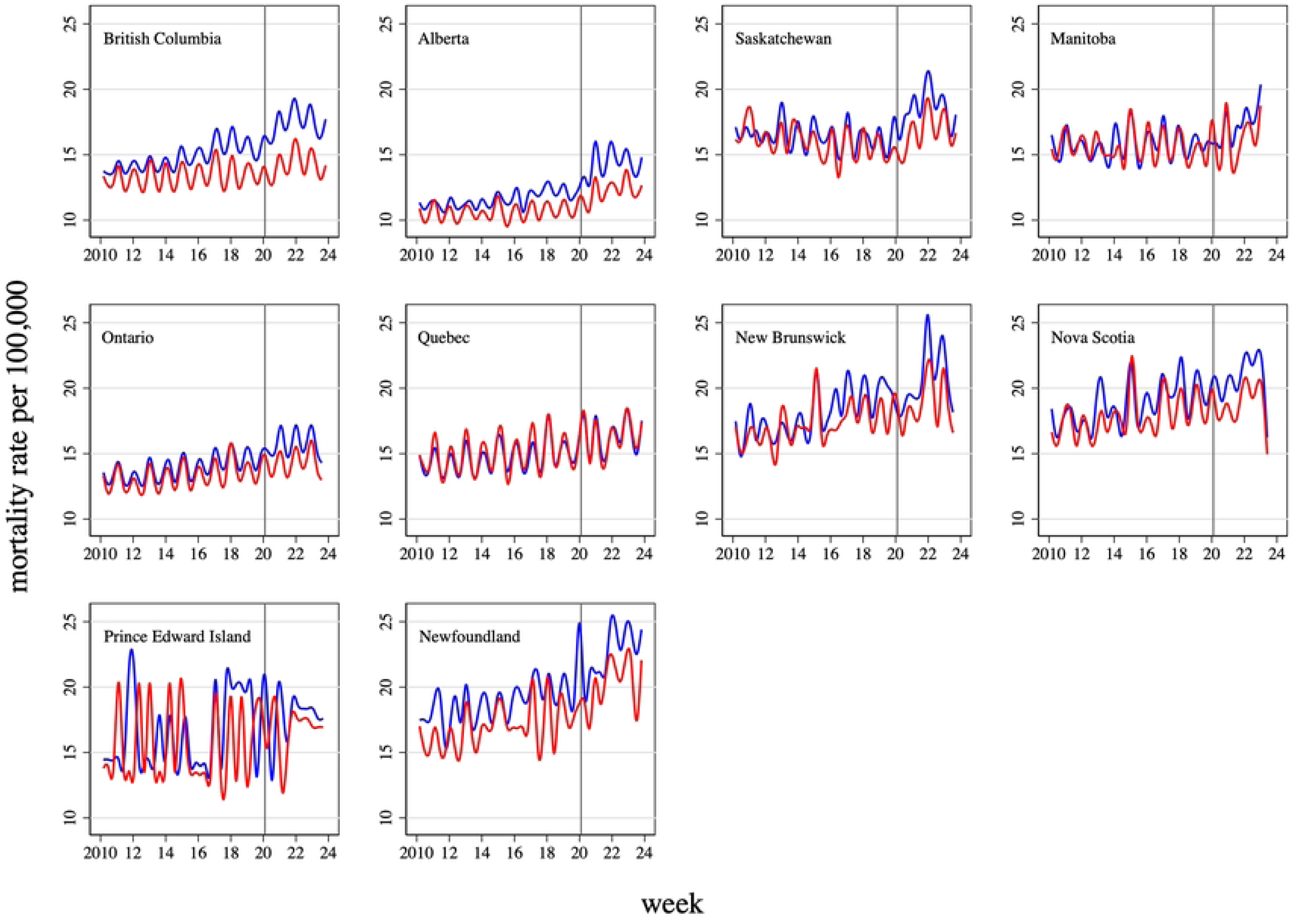
Median spline curves fit to province and sex stratified data on all-cause mortality rates per 100,000 population, by week, 2010-2023. Data source: Statistics Canada. Table 13-10-0768-01 Provisional weekly death counts, by age group and sex. Population data: Statistics Canada. Table 17-10-0005-01 Population estimates on July 1, by age and gender. Blue line: fitted spline for males. Red line: fitted spline for females. Vertical line indicates the week that WHO declared the global COVID-19 pandemic.

Figs 11 and 12 show trends in life expectancy at birth and at 65, respectively, providing insights into the sex and age differentiated mortality impacts of the COVID pandemic. Alberta and Saskatchewan experienced the largest reductions in life expectancy at birth (Fig 11). The estimated reductions, illustrated in Appendix 4 and reported by year in S1 Table and as 3-year averages in Table 1, are also generally larger for males than for females. For instance, in Alberta, the average annual reduction in life expectancy at birth during the three pandemic years was 1.8 years for males but 1.2 years for females. The reduction in life expectancy by year in Fig 11 generally matches the spikes in COVID-attributed observed earlier. For instance, the surge in COVID-attributed mortality in New Brunswick observed after the easing of controls in 2022 (Fig 6) is reflected in the reduction in life expectancy for that year. There were exceptions, however. The reductions in life expectancy in Newfoundland were not observed in the COVID-attributed mortality data. There was also marked variation across provinces in the impacts of COVID on life expectancy at 65 (Fig 12 and S1 Table). As was the case for life expectancy at birth, the declines in life expectancy at 65 were largest in Saskatchewan.

**Fig 11.**
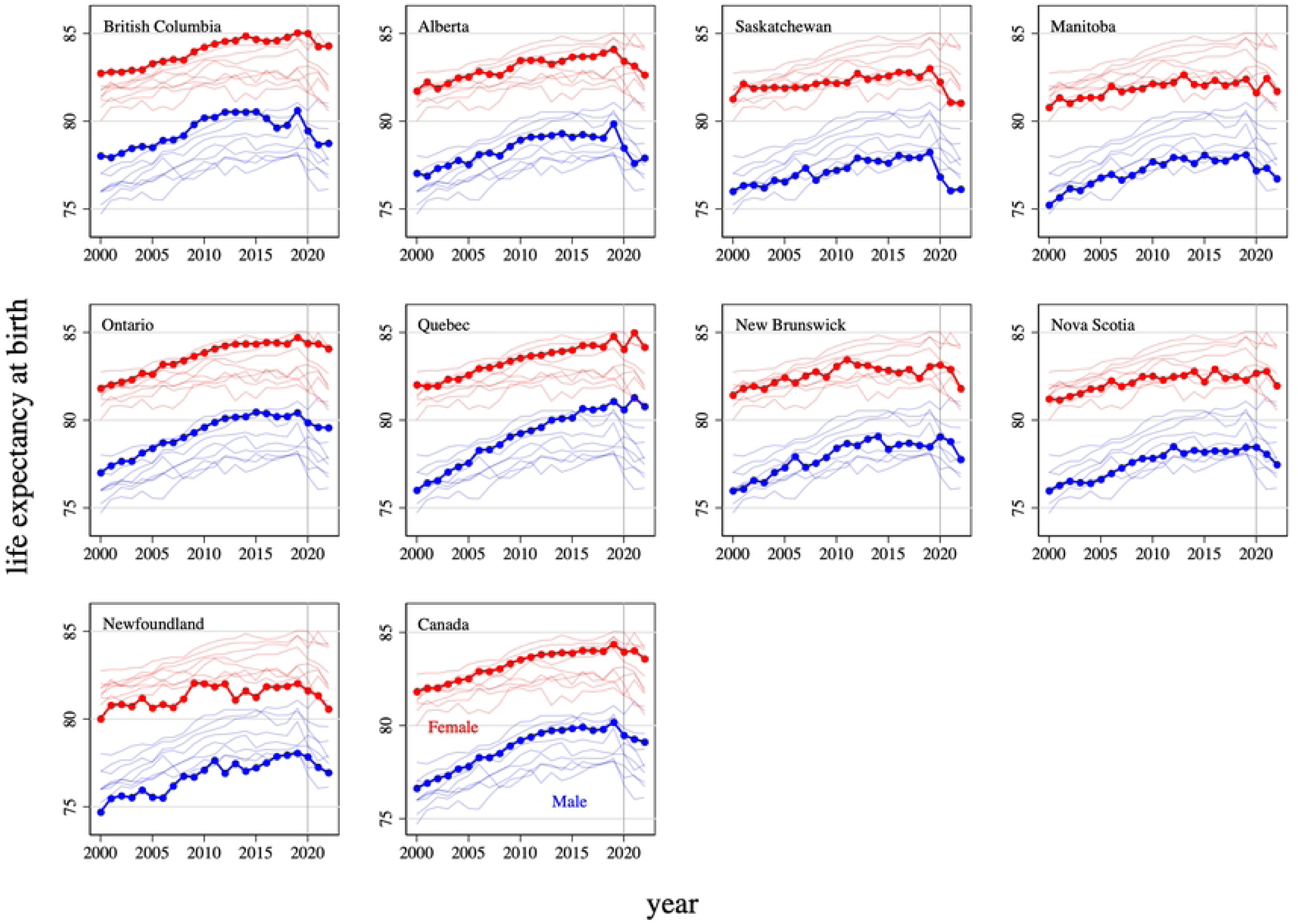
Life expectancy at birth, by sex, province and year, 2000-2022. Data source: Statistics Canada. Table 13-10-0837-01 Life expectancy and other elements of the complete life table, single-year estimates, Canada, all provinces except Prince Edward Island. Blue line: males. Red line: females. Vertical line indicates the year that WHO declared the global COVID-19 pandemic. Each graph plots life expectancy for all 9 provinces, but bolds the life expectancy for just one province or for Canada.

**Fig 12.**
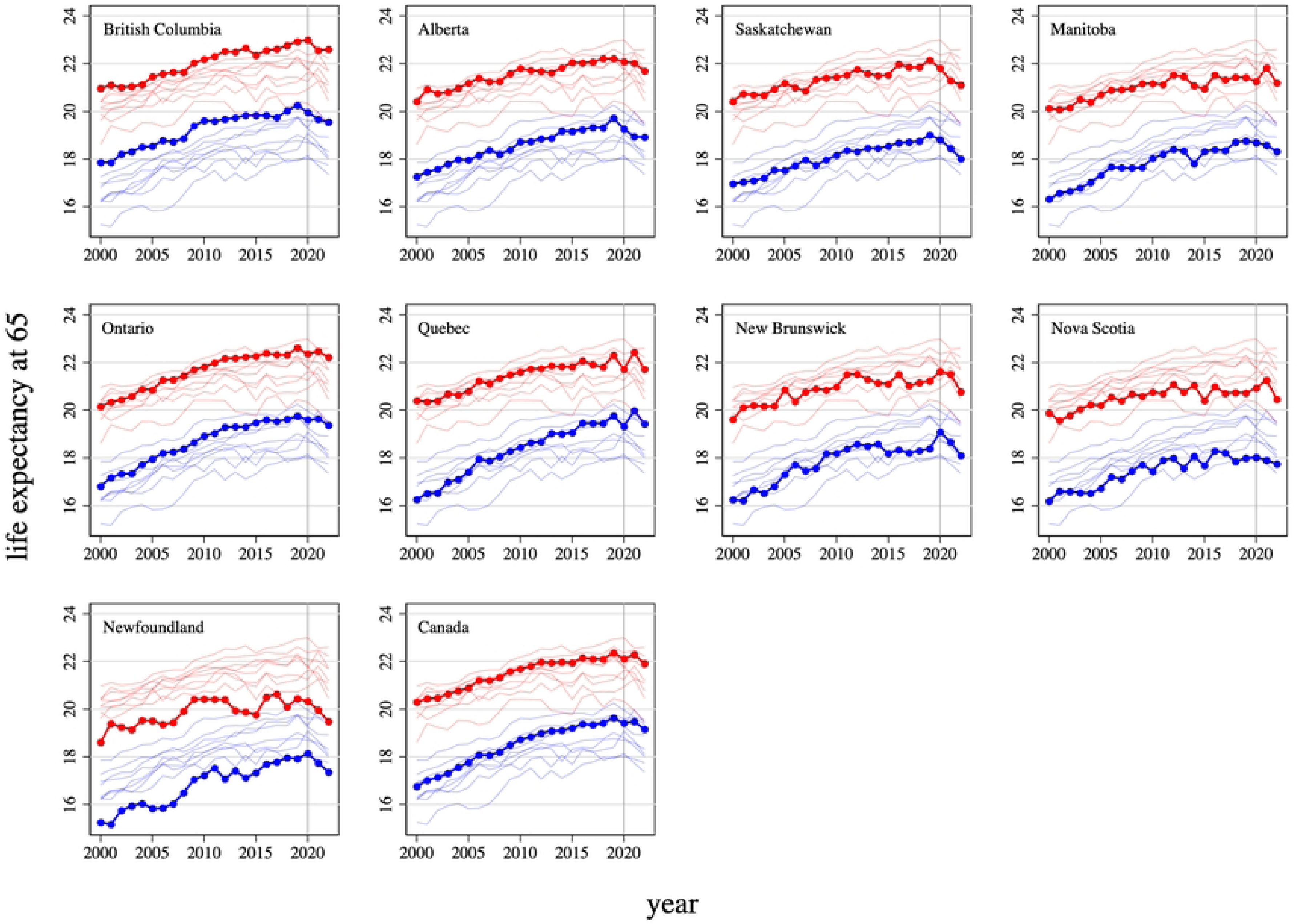
Life expectancy at 65, by sex, province and year, 2000-2022. Data source: Statistics Canada. Table 13-10-0837-01 Life expectancy and other elements of the complete life table, single-year estimates, Canada, all provinces except Prince Edward Island. Blue line: males. Red line: females. Vertical line indicates the year that WHO declared the global COVID-19 pandemic. Each graph plots life expectancy for all 9 provinces, but bolds the life expectancy for just one province or for Canada.

### Interpretation

Razak and colleagues found that during the first two years of the pandemic (2020 and 2021), compared to its peers, Canada used relatively stringent public health measures, performed better than most in terms of COVID vaccination rates and excess mortality, but experienced the largest percentage declines in per capita GDP. [1]

This paper uses longer term data to provide insights into provincial level variation in pandemic responses and outcomes. The provinces varied in their use of the most stringent public health measures. Compared to Alberta and Saskatchewan, Ontario and Quebec used stay-at-home orders, strict limits on gatherings and other restrictive public health measures for longer periods throughout the pandemic. The Atlantic provinces received the lowest volume of international arrivals in the months prior to the official start of the pandemic. These provinces were able to contain the pandemic using strict border controls and short, judiciously applied lockdowns. This perhaps explains why this region experienced the smallest GDP reductions. The uptake of the primary COVID vaccinations varied from 76% (in Alberta) to 92% (in Newfoundland). Uptake of the booster vaccinations during 2022 and 2023 was much lower than for the primary series; there was also greater interprovincial variation. Excess mortality and economic outcomes differed markedly by province. For example, average annual estimated changes in female life expectancy at 65 during the years 2020-2022 varied from a small (0.1 year) increase in New Brunswick to a 0.9 year decrease in Saskatchewan.

While this study does not formally estimate the impacts of provincial response stringency and COVID vaccine uptake on mortality and other outcomes, it does provide suggestive evidence. The higher excess mortality among males than females – also observed outside Canada [30] – could be due to the lower male vaccination rates.

Uptake of the primary COVID vaccinations and the booster vaccinations among males were, respectively, about 2-3 percentage points and 5-6 percentage points lower than for females. Again, these findings are merely suggestive. These mortality differences could also be due to sex differences in immune responses to SARS-CoV-2 infection or compliance with public health restrictions. [31]

The data presented here also suggests that the most stringent provincial infection control measures were effective. The reason is that surges in COVID-attributed mortality abated after the imposition of these measures. Moreover, outside of Ontario and Quebec, rates of excess mortality tended to be highest after most stringent infection control measures were eased in the first half of 2022. At the national level, the lowest pandemic-era life expectancy values, both at birth and at 65, were observed in 2022.

This study also finds the highest excess mortality in Alberta and Saskatchewan, provinces with the lowest response stringency levels after the first wave of the pandemic. Compared to Alberta and Saskatchewan, Ontario and Quebec used the most restrictive public health measures for longer periods throughout the pandemic. Ontario and Quebec also experienced much lower excess mortality. Again, other factors could be at play: Alberta and Saskatchewan also had the lowest vaccination uptake (both primary and booster doses).

This study corroborates findings reported elsewhere about apparent underestimation of COVID mortality rates reported by public health authorities in some provinces. [6] For instance, the surge in excess mortality in Newfoundland in 2022 was not reflected in the COVID-attributed mortality data reported by the provincial health authorities.

Research on the impacts of provincial infection mitigation strategies and vaccine uptake, some of which is already appearing in the literature, [32,33] will help to inform government responses to future pandemics. This research enterprise should use methods that overcome the limitations of the excess mortality approach used in this study. In some provinces, the variability in GDP and life expectancy in the years before the pandemic makes it challenging to predict what outcomes would have been in the absence of the pandemic. The excess mortality approach also cannot distinguish the causes of pandemic-era excess mortality and whether these causes of death were influenced by the pandemic. For instance, it is unclear to what extent excess mortality is due to opioid toxicity and whether these tragic deaths were in part caused by the pandemic or would have occurred anyways.

## Conclusion

Canada’s provinces used markedly different approaches to controlling the spread of SARS-CoV-2 infection; rates of primary COVID vaccine uptake also varied across provinces. The provinces also experienced markedly different excess mortality and economic outcomes. These outcomes appear to be related to provincial responses. To better prepare for future pandemics, additional research should help elucidate the mechanisms linking these province-level responses to outcomes.

## Data Availability

All data used in this study are publicly available online. If the study is accepted for publication in PLOS ONE, the data will also be published in Borealis, a public data repository.

https://doi.org/10.25318/1310083701-eng

https://doi.org/10.25318/1710000501-eng

https://doi.org/10.25318/1310076801-eng

https://doi.org/10.25318/1310078401-eng

https://doi.org/10.25318/3610022201-eng

https://doi.org/10.34989/SAN-2021-1

https://health-infobase.canada.ca/src/data/covidLive/covid19-download.csv

https://health-infobase.canada.ca/covid-19/vaccination-coverage/

https://odesi.ca/en/details?id=/odesi/doi__10-5683_SP3_LEVP0M.xml

https://doi.org/10.25318/2410005301-eng

## Acknowledgments

The author thanks seminar participants at the Institute for Pandemics (University of Toronto) and especially Mehdi Ammi, Nelson Lee, Alexander Meek, and Sharifa Nasreen for helpful comments.

## Supporting information

**S1 Fig. Real per capita gross domestic product, by province and year, 2010-2022, and projected real per capita gross domestic product during 2020-2022 based on 2015-2019 trend line.** Data source: GDP: Statistics Canada. Table 36-10-0222-01 Gross domestic product, expenditure-based, provincial and territorial, annual. Population data: Statistics Canada. Table 17-10-0005-01 Population estimates on July 1, by age and gender. Green line: 2015-2019 trend. Vertical line indicates the year that WHO declared the global COVID-19 pandemic. Each graph plots real per capita GDP for each province, but bolds the real per capita GDP for just one province.

**S2 Fig. Life expectancy at birth, by sex, province and year, 2010-2022, and projected life expectancy at birth during 2020-2022 based on 2015-2019 trend line.** Data source: Statistics Canada. Table 13-10-0837-01 Life expectancy and other elements of the complete life table, single-year estimates, Canada, all provinces except Prince Edward Island. Blue line: males. Red line: females. Green line: 2015-2019 trend. Vertical line indicates the year that WHO declared the global COVID-19 pandemic. Each graph plots life expectancy for each province, but bolds life expectancy for just one province.

**S3 Fig. Life expectancy at 65, by sex, province and year, 2010-2022, and projected life expectancy at 65 during 2020-2022 based on 2015-2019 trend line.** Data source: Statistics Canada. Table 13-10-0837-01 Life expectancy and other elements of the complete life table, single-year estimates, Canada, all provinces except Prince Edward Island. Blue line: males. Red line: females. Green line: 2015-2019 trend. Vertical line indicates the year that WHO declared the global COVID-19 pandemic. Each graph plots life expectancy for each province, but bolds life expectancy for just one province.

**S1 Table. Estimated post-pandemic changes in outcomes along with 95% confidence intervals, by province, year, 2020-2022, and outcome (real per capita gross domestic product, life expectancy at birth, life expectancy at 65).** Note: the province-year estimates were obtained by estimating, using annual data from 2015-2022 for each province, a linear regression of the outcome variable on a linear time trend and separate indicator variables for 2020, 2021 and 2022 years.

